# Sequencing and health data resource of children of African ancestry

**DOI:** 10.1101/2025.03.22.25324419

**Authors:** Leah C. Kottyan, Scott Richards, Morgan E. Tracy, Lucinda P. Lawson, Isabelle C. Kottyan, Annika Curwin, Beth Cobb, Steve Esslinger, Margaret Gerwe, James Morgan, Alka Chandel, Leksi Travitz, Yongbo Huang, Catherine Black, Agboade Sobowale, Tinuke Akintobi, Monica Mitchell, Andrew F. Beck, Ndidi Unaka, Michael Seid, Sonja Fairbanks, Michelle Adams, Tesfaye Mersha, Bahram Namjou, Michael W. Pauciulo, Jeffrey R. Strawn, Robert T. Ammerman, Daniel Santel, John Pestian, Tracy Glauser, Cynthia A. Prows, Lisa J. Martin, Louis Muglia, John B. Harley, Iouri Chepelev, Kenneth M. Kaufman

**Author notes:** Corresponding authors: Leah Kottyan; Kenneth Kaufman.

## Abstract

**Purpose:** Individuals who self-report as Black or African American are historically underrepresented in genome-wide studies of disease risk, a disparity particularly evident in pediatric disease research. To address this gap, Cincinnati Children’s Hospital Medical Center (CCHMC) established a biorepository and developed a comprehensive DNA sequencing resource including 15,684 individuals who self-identified as African American or Black and received care at CCHMC.

**Methods:** Participants were enrolled through the CCHMC Discover Together Biobank and sequenced. Admixture analyses confirmed the genetic ancestry of the cohort, which was then linked to electronic medical records.

**Results:** High-quality genome-wide genotypes from common variants accompanied by medical record-sourced data are available through the Genomic Information Commons. This dataset performs well in genetic studies. Specifically, we replicated known associations in sickle cell disease (*HBB*, HGNC:4827, p = 4.05 × 10⁻¹LL), anxiety (*PLAAT3*, HGNC:17825, p = 6.93 × 10⁻L), and asthma (*PCDH15*, HGNC:14674, p = 5.6 × 10⁻¹L), while also identifying novel loci associated with anxiety, asthma, and asthma severity.

**Conclusion:** We present the acquisition and quality of genetic and disease-associated data and present an analytical framework for using this resource. In partnership with a community advisory council, we have co-developed a valuable framework for data use and future research.

## Introduction

Individuals of self-reported African ancestry have historically been underrepresented in genetic studies of diseases, reflecting broad disparities in genetic research and clinical medicine^1, 2^. This underrepresentation stems from systemic inequities in research funding, recruitment practices, and access to healthcare, which together limit the inclusion of diverse populations in study cohorts^1–4^. As a result, most genomic data have been derived from individuals of European ancestry, leading to significant gaps in understanding the genetic underpinnings of diseases affecting Black or African American adults and children^5, 6^. This lack of diversity undermines the ability to identify ancestry-specific genetic variants, which reduces the applicability of genomic findings, precision medicine interventions, and risk prediction models for certain populations^1, 7, 8^. Addressing ancestry-based disparities in research is critical for ensuring equitable advances in healthcare and bridging gaps in disease prevention and treatment outcomes for underrepresented groups^1^.

The underrepresentation of individuals of African ancestry in genome-wide studies of disease risk has been a persistent issue, particularly in the context of pediatric diseases^2, 9^. This lack of representation not only limits our understanding of disease etiology across ancestries, but hampers the development of effective, equitable healthcare solutions^1, 3^. In 2024, 19.8% of the patients who received care at CCHMC and 15.6% of patients enrolled in the Discover Together Biobank with a DNA sample available identified as Black or African American. To address this critical gap, CCHMC has undertaken a significant initiative to create a comprehensive sequencing resource focused on broadly expanding opportunities to identify the genetic basis of health and disease in children of African ancestry.

The primary objective of this study was to acquire high-quality genetic data from participants in the Discover Together Biobank who self-identified as Black or African American and had accompanying electronic medical records. To achieve this goal, we employed a low-pass sequencing strategy^10–12^, which yielded an average genomic coverage of around 3X (i.e. meaning that the average number of reads per base is ∼3). The low pass approach enabled us to significantly broaden the scope of our study compared to what would have been feasible with standard 30X read depth genome sequencing.

To demonstrate the utility of this resource, our secondary objective was to conduct GWAS analyses for three diverse human disorders; sickle cell disease (OMIM #603903), anxiety disorder (OMIM #607834), and asthma (OMIM #600807). Sickle cell disease is a severe inherited blood disorder that predominantly affects Black and African American children in the United States, with an estimated prevalence of approximately 1 in 365 African American births in this population^13, 14^. It is almost always caused by at least one hemoglobin S allele in combination with a second pathogenic variant in the *HBB* gene (HGNC:4827) resulting in the production of abnormal hemoglobin S^14, 15^. When deoxygenated, hemoglobin S polymerizes, leading to the characteristic sickling of red blood cells, increased hemolysis, and vaso-occlusive events^16^. Clinically, sickle cell disease manifests with episodes of severe pain (vaso-occlusive crises), anemia, increased susceptibility to infections, and progressive organ damage, significantly impacting quality of life and life expectancy^13^.

Anxiety disorders are among the most common mental health conditions in children, with prevalence estimates ranging from 7% to 10%, though rates may be higher due to underdiagnosis in some populations^17^. Black and African American children experience anxiety at similar or higher rates than their White peers, yet they are less likely to receive a diagnosis or access appropriate mental health care^18, 19^. While genetic studies have identified variants associated with anxiety risk, there has been limited research specifically examining genetic contributors in African American youth, leading to gaps in understanding potential ancestry-specific risk factors.

Asthma is one of the most common chronic diseases in children, with a disproportionate burden among Black and African American children, who experience higher prevalence, increased severity, and greater rates of hospitalization and mortality compared to other racial and ethnic groups^20, 21^. In the United States, Black children have nearly twice the prevalence of asthma as White children, with environmental, socioeconomic, and genetic factors contributing to this disparity^22, 23^. Genetic studies have identified multiple loci associated with asthma risk, including variants in genes such as *TSLP*^24^ (HGNC:30743), *GSDMB*^25–27^ (HGNC:55876), and *IL1RL1*^28^ (HGNC:5998), though much of the research has been conducted in populations of European ancestry, limiting insights into genetic contributors in African American pediatric cohorts^29^.

Herein, we present a CCHMC-based biobank resource, which encompasses deep genetic and phenotypic data from 15,684 individuals who self-identified as African American or Black and received care at CCHMC. We detail the acquisition and quality of the genetic and health data contained in this resource, and we also recommend an analytical framework for using these data alongside various analyses that demonstrate the power and potential of this unique resource. Importantly, this initiative has been developed in partnership with a community advisory council, ensuring that the framework for data use and future research is aligned with community needs and perspectives (see accompanying perspectives piece).

## Methods

### Collaboration with a community advisory committee

Meeting every other month over the course of two years, the committee facilitated discussions focused on ensuring that data and biological samples were kept safe while promoting transparency in research practices. A key focus was increasing the representation of historically underrepresented populations, particularly Black and African American children, in genomic studies to address longstanding disparities in research participation. The committee worked collaboratively to develop and review community engagement strategies, identify effective methods of communication, and strengthen relationships between community-engaged research and partners. The insights from this group directly influenced how we presented data from the cohort and shaped our decision to prioritize genetic studies of asthma and anxiety, conditions that disproportionately impact Black and African American children (**See co-submitted Perspective article submitted by members of this advisory committee**).

### Discover Together Biobank

The CCHMC Discover Together Biobank is an actively-enrolling, multifaceted initiative that collects biospecimens with associated phenotypic and genotypic data while combining clinical research protocols with appropriate governance and accession processes. Discover Together Biobank serves as a key piece of infrastructure to aid the research of CCHMC investigators and currently contains samples and electronic medical record data from over 100,000 participants. Discover Together Biobank is constructed from a patient-based participant population that spans multiple institutional biobanking consent periods at CCHMC.

Participants for this study were consented to the CCHMC IRB approved study during a consent period as part of our “Better Outcomes for Children” cohort, which used broad consent to enroll CCHMC patients to ascertain clinical residual blood samples for approved research use. The Better Outcomes for Children consent allows for use of DNA and associated medical record data for research, including genomics. The self-reported race information was identified from Cincinnati Children’s electronic medical record system of Epic using Epic Clarity from the ZC_PATIENT_RACE.NAME field. Registrars at Cincinnati Children’s Hospital collect race information from patient families at the time of patient care.

### Deposition of Data in the Genetic Information Commons

Data from this sequencing resource is available through the Genomic Information Commons (GIC). The GIC is a research initiative allowing researchers to collaborate across multiple academic medical centers to share consented patient genomic data for large-scale research projects aimed at discovery of genetic mechanisms of health and disease and improving patient health outcomes (https://www.genomicinformationcommons.org/)^30^. The GIC facilitates collaborative research by enabling sharing of genomic data, phenotypic information, and biospecimen metadata across participating institutions. For more information on data requests, please contact.

#### DNA preparation and sequencing

See Supplemental Note (**Supplemental Methods**). **Supplemental Dataset 1** contains information about the chromosomal position (CHR), variant names (SNP), reference (A1) and alterative alleles (A2), and the non-missing allele count (NCHROBS) for each variant in the dataset.

### Principal component analysis and ancestry assessment

Principal component analysis was performed using the software package Plink2 (version 2.0-161017). An LD pruned data set was generated with Plink2 (independent-pairwise 50kb window, 5 step, 0.8 r^2^ threshold and minor allele frequency between 0.15 and 0.4). The first 10 principal components were approximated using Plink2. Admixture for each sample was calculated using the software program admixture version 1.3.0 assuming 4 populations^31^. 155 individuals had less than 2% African admixture and were removed from this cohort. **Supplemental Dataset 2** contains Principal Component analysis and ancestry information for each sample.

### Assessment of relatedness between samples

Family relationships were determined using the KING software package (version 2.1.6)^32^. **Supplemental Dataset 3** provides relatedness information for this cohort.

### Genome-wide association studies

Study cohorts were generated by identifying cases in the biorepository samples with matching ICD 10 codes. Only one case per family was used for analysis. All families with an affected sample were dropped as a source for potential control samples. Cases were matched to controls using the R package PCAmatchR (5^th^ release) based on the first three principal components^33^. Cases were matched to controls with a match distance less than 0.1 as determined by PCAmatchR. The number of controls per case was dependent on the total number of cases and available potential controls.

Quality control of genotype data was performed on each cohort. Variants with a case minor allele frequency less than 1% or a variant call rate below 85% were dropped. In addition, any variant with a control HWE p-value below 0.0001 was dropped. All QC measurements and association analyses were performed with the Plink1.90b6.26^34^. After identifying controls that were matched to cases based on admixture and removing first and second-degree relatives, genome wide association analyses were performed in PLINK1.9 using a logistic regression that adjusted for admixture estimates and genomic inflation.

Interactive Manhatton plots are available for the association studies presented in this manuscript:

Sickle cell disease vs controls:

https://my.locuszoom.org/gwas/378516/?token=788c8c35d21348b996cd25c638d4ebc3

Anxiety vs controls:

https://my.locuszoom.org/gwas/852976/?token=57f16f9e8123412192f061c71246f89c

Asthma vs controls:

https://my.locuszoom.org/gwas/407471/?token=ab55b1868cb141ac979a84b3e6859c10

Severe versus controlled asthma: https://my.locuszoom.org/gwas/48795/?token=6823af24c2f94f89af20ce3a20b0b6ca

See **Supplemental Note** for a suggested protocol for case - control studies using this resource.

### Partnerships to connect expert clinical phenotyping with high quality genetic data: acute asthma exacerbation

We partnered with clinical experts in the Asthma Learning Health System,^21^ an asthma-focused learning network at Cincinnati Children’s, to identify carefully phenotyped groups and perform clinically directed analyses. The Asthma Learning Health System aims to advance equitable asthma care, with the goal that all children with asthma can live their best lives. The network’s clinical team identified children treated at Cincinnati Children’s to manage a diagnosis of asthma, further identifying two key groups: 99 participants who required acute care in the emergency department for asthma and 393 participants who managed their condition without needing acute interventions during the study period. Leveraging genomic data from this sequencing resource, we conducted a genome-wide association study (GWAS) to investigate genetic factors contributing to asthma severity and healthcare utilization.

## Results

A Community Advisory Committee played a critical role in guiding the way we developed this sequencing research. Including Community Advisory Committee perspectives was and remains critical as we strive for maximally ethical and inclusive conduct of pediatric genetic research, particularly in relation to biorepositories and data security (**see co-submitted Perspectives article authored by members of advisory committee**).

The Discover Together Biobank supplied 16,853 samples for this project. These samples were processed in batches of 960 for barcoding and pooling into a sequencing library. Following sequencing, the samples were demultiplexed based on sample-specific barcodes and aligned to the human GRCh38 reference genome. Imputation was performed to enhance genotype accuracy and coverage using the Sequencing To Imputation Through Constructing Haplotypes (STITCH) approach, which does not require a reference panel^35^. To provide more genetic diversity in each imputation assessment, down-sampled sequence data from the 1000 genomes project^36^ were included in each imputation batch. Data quality was assessed based on sample and variant-level metrics, as well as the concordance of genotypes with available 30x whole genome sequence data. Samples were removed if the sample call rate was less than 85% or the reported gender and imputed sex were discordant. Duplicated samples were removed from the dataset unless the records indicated that the samples were obtained from twins who were independently consented into the Discover Together Biobank.

DNA from control samples with established genome sequences were included to support robust quality assessments focused on the concordance of heterozygous variants (see **Supplemental Methods** and **Supplemental Figures 1 and 2**). The final dataset contains high quality sequence data for 15,684 samples that correspond to consented patients with electronic medical record data at Cincinnati Children’s. In this sequencing resource, 53.5% of the samples were from females (**Figure 1A**). The average genomic coverage was 2.66 (median: 2.60, interquartile range: 1.12) (**Figure 1B**). The mean age of enrollment was 9.68 (with a standard deviation of 7.33) years old, and the average current age of participants is 19.77 years. Samples had an average of 3,155,811 called variants with allele frequencies greater than 1%, with a range from 2,542,625 to 3,417,031 (**Figure 1C**). The average per variant call rate was 93.2%, ranging from 84.4%-96.7% (**Figure 1D**). The allele frequency of variants in this resource are highly correlated (R^2^ = 0.988, Person p<0.0001) with those in reported as the African alternative allele frequency in the gnomaAD database (**Supplemental Figure 3**). **Supplemental Dataset 1** provides a list of all variants in this resource.

**Figure 1.**
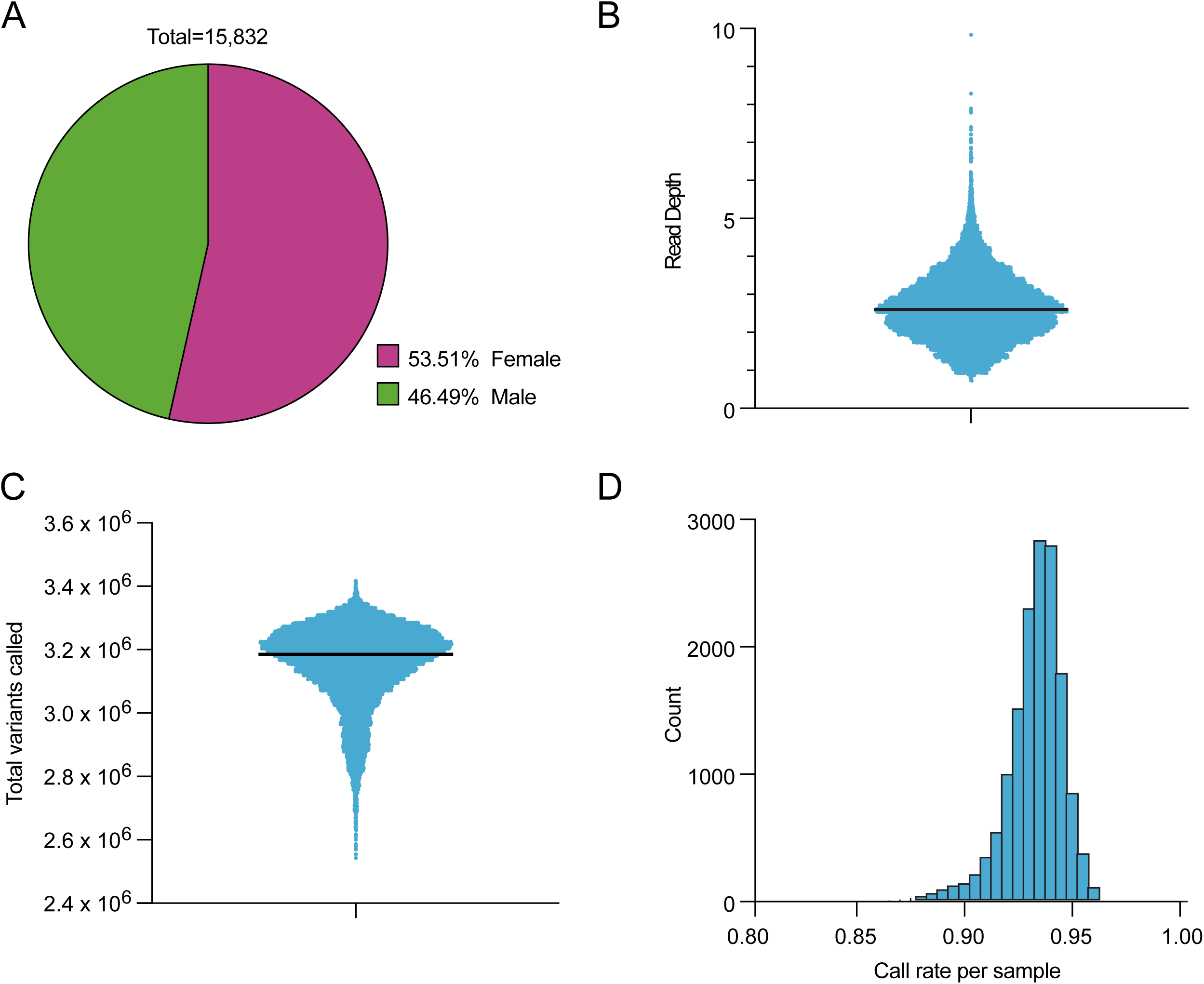
Genotype data of the cohort. A. Distribution of female and male participants. B. Genomic coverage with a line indicating the median. C. Distribution of total genetic variants called with a line indicating the median. D. Per participant call rate of variants included in the final dataset.

**Figure 2.**
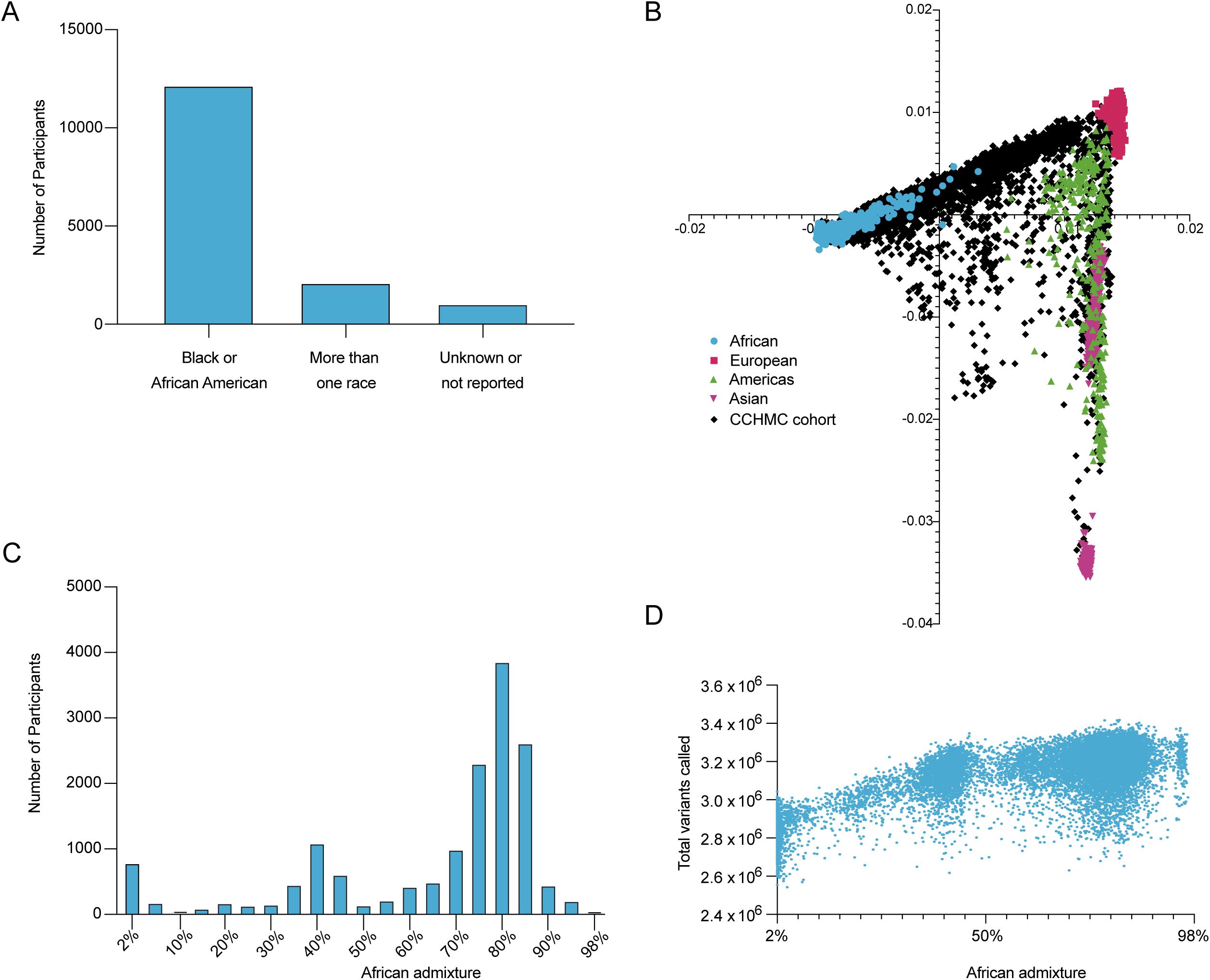
Ancestry of children in the cohort. A. Self-reported race as collected in the electronic medical record for each participant. B. Principal component analysis of each participant (black dots) in the context of reference individuals from the 1000 Genomes Project, whose ancestry is indicated by color (see legend). C. Distribution of African admixture across individuals in the cohort. D. The number of variants called per sample (dots) in relationship to the genomic proportion of African admixture. CCHMC: Cincinnati Children’s Hospital Medical Center.

A total of 77.8% of the participants in this study self-identified as Black or African American to the hospital registrar as documented in their corresponding electronic medical record data (See Methods), with the remaining individuals identifying as more than one race and/or ethnicity or did not provide data (**Figure 2A**). We performed principal component and admixture analysis to identify genetic ancestry, using 1000 genomes data as a reference (**Figure 2 B and C**). As expected, most participants in this study had 75-85% African admixture. Strikingly, there was a large range of African admixture in this cohort, ranging from 2-98% (**Figure 2C** and **Supplemental Dataset 2**). The number of non-reference (alternative) alleles called within each sample was strongly correlated with the proportion of African ancestry (**Figure 2D**), which is consistent with the enhanced genetic diversity of the populations from Africa relative to other global populations ^37, 38^.

As expected from a pediatric biobank-based study, there was substantial relatedness between participants in the cohort (**Table 1**). A total of 1,050 siblings were identified in addition to 41 monozygotic twin pairs. Also, 98 parent-offspring pairs were identified, providing opportunities to study genetic heritability in the context of specific health outcomes; 134 2^nd^ degree relatives were also identified (**Table 1**). A relatedness matrix was developed to support additional studies using this cohort (**Supplemental Dataset 3**). Altogether, these results highlight opportunities to study health outcomes within families and underscore the importance of accounting for relatives in case-control based studies within this resource.

### Clinical characteristics of the cohort

Electronic medical records linked to the participants in this study include 10,472 distinct ICD 10 codes, with the top 20 codes listed in (**Table 2**). The most common codes include those for infections, common symptoms of childhood illnesses, gastrointestinal complaints, attention-deficit hyperactivity disorder, obesity, and dermatitis. Other top ICD 10 codes were associated with routine medical care of children including encounters for immunizations. Current Procedure Terminology (CPT) codes are used to report medical, surgical, and diagnostic procedures and services, while Healthcare Common Procedure Coding System (HCPCS) codes report medical procedures, supplies, and services that are not included in CPT. In this cohort, 18,708 distinct CPT and HCPCS codes were reported. The top reported codes in this cohort were for medical visits (ranging from routine visits for established patients to complex emergent encounters), venipuncture (with associated blood tests), vaccination, urinalysis, tonsillectomy with adenoidectomy, and spirometry (**Table 3**). Of the 11,717 distinct medication codes used in this cohort, the most common were analgesics, intravenous saline, antiemetics, albuterol, antibiotics, and steroids (**Table 4**). Although participants are largely from Metropolitan Cincinnati, the area surrounding CCHMC, home addresses were reported across most of the United States (**Figure 3**). Because street addresses and zip codes are available from each medical encounter of a participant within the CCHMC’s system, studies integrating genetic data with environmental data gleaned from geocoding are possible.

**Figure 3.**
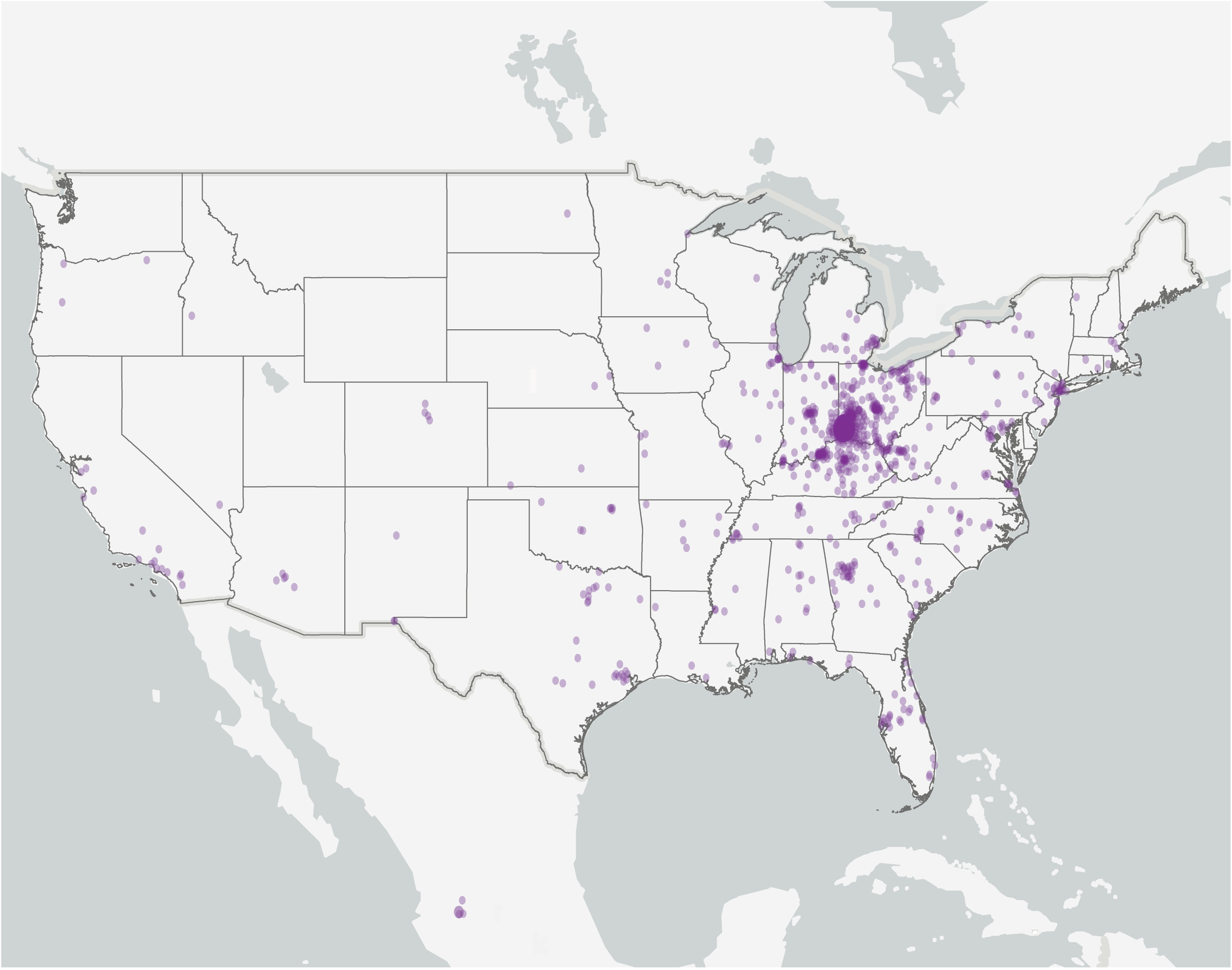
Geographic distribution of children in this cohort. Each dot represents a zip code with at least one participant in this study. Dots are sized and have transparency adjusted based on the number of participants who lived in the corresponding zip code. All zip codes were geocoded and plotted with Esri ArcGIS Pro 3.4. Map sources: Esri, TomTom, Garmin, FAO, NOAA, USGS, EPA, USFWS.

### Exemplary case control genome wide association studies using this cohort

We sought to establish the utility of this resource through exemplary genome-wide association studies. The phenotypes we focused upon were identified in collaboration with our community advisory committee. The number of cases and controls, information about the sex and African admixture of cases and controls, as well as the genomic inflation for each association study presented are provided in **Table 5**.

#### Sickle-cell Disorder

In our cohort, 754 individuals had the D57 ICD-10 code (coding for sickle-cell disorders) and were identified as “cases”. Using the framework presented in **Supplemental Figure 4**, 4,484 controls were identified. Because this sequencing resource includes common variants, this analysis was intended to identify modifiers of sickle-cell disorder. Rs334 is a variant in the β-globin gene (*HBB*), which in homozygous form causes sickle cell disease. Because rs334 has two alternative alleles, its genotype was not called for this dataset. The most significant genetic association identified in this analysis consisted of a group of variants in a genetic haplotype tagged by rs4426157 (p-value = 4.05×10^−148^; odds ratio = 4.38) (**Figure 4A** and **Supplemental Dataset 4**), which is located six kilobases downstream of *HBB*. rs334 and rs4426157 are in moderate linkage disequilibrium, and a conditional logistic regression analysis accounting for the genotype at rs4426157 markedly reduced genetic association in the region (**Supplemental Dataset 4**). Variants in this haplotype have also been identified in independent cohorts assessing hemolysis in sickle cell anemia (10^−10^ < p-values <10^−29^)^39^ and thromboembolic events in sickle cell disease (10^−6^ < p-values <10^−8^)^40^.

**Figure 4.**
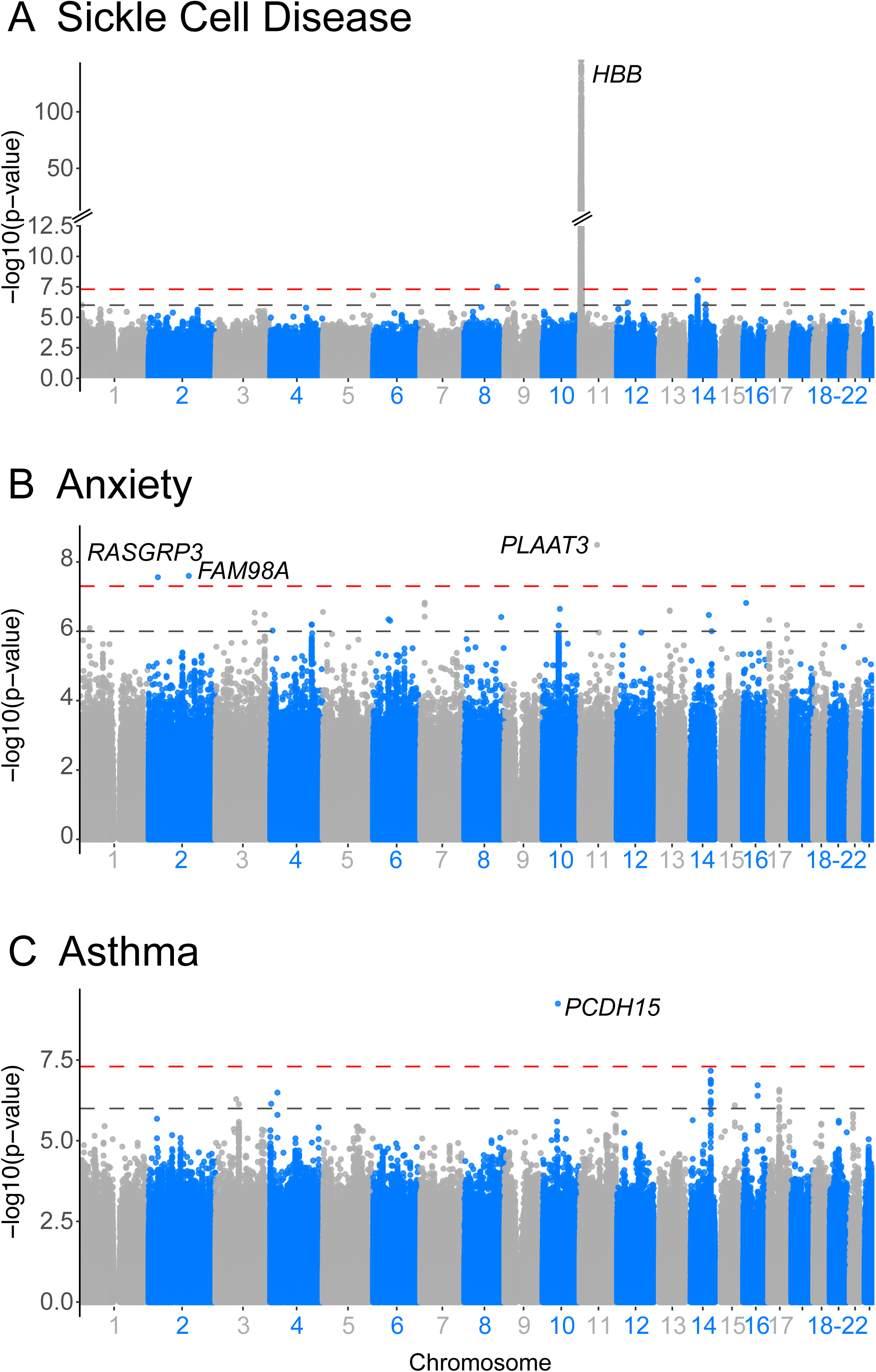
Exemplary case-control genome-wide associations studies using this cohort. Genome-wide association studies were performed using the framework presented in **Supplemental Figure 4** for sickle cell disorders (note break on y-axis) (A), anxiety (B), and asthma (C). For each study, the negative log p-value of a logistic regression analysis that accounts for the first three principal components is shown. Genomic inflation for these analyses were 1.033, 1.048, and 1.042 for sickle cell disease, anxiety, and asthma, respectively.

#### Anxiety disorders

Using a clinical phenotyping algorithm developed by CCHMC investigators, we identified 3,733 participants with anxiety in this cohort. After identifying 7,341 controls, we performed an association study and found three variants with genome-wide association (**Figure 4B** and **Supplemental Dataset 4**): rs78667939 near *PLAAT3* (p-value = 6.93×10^−9^; odds ratio = 1.55), rs77200838 in an intergenic region (p-value = 2.75×10^−8^; odds ratio of risk allele = 1.58), and rs181943174 near *FAM98A* (HGNC:24520) and *RASGRP3* (HGNC:14545) (p-value = 2.12×10^−8^; odds ratio = 2.09). This study is the first to report these three novel genome-wide associated anxiety risk loci.

#### Asthma

After identifying 4,303 cases with electronic medical record codes of ICD-10 J45 and 8,342 principal component-matched controls, we performed a genome wide association study. We identified one locus with a variant exceeding genome-wide significance: rs185118029 near *PCDH15* (HGNC:14674, 5.6×10^−10^; odds ratio = 1.71) (**Figure 4C** and **Supplemental Dataset 4**). Variants at the *PCDH15* locus have not previously been identified for asthma. Additionally, numerous risk loci from our analysis with p-values ranging from 10^−4^ to 10^−7^ included variants that were previously reported as increasing asthma risk in independent studies. These included replication of known asthma risk loci at *GSDMB*^26, 27^ (HGNC:23690), *IL1RL1*^41, 42^ *(HGNC:5998)*, *TSLP*^43^ (HGNC:2539), *RAD50*^44^ (HGNC:9816), *D2HGDH*^41, 42, 44–52^ (HGNC:28358), *CDHR3*^44^ (HGNC:26308), and *PRDM11*^46^ (HGNC:13996) (**Supplementary Dataset 5**).

### Asthma Learning Health System acute asthma GWAS results

The Asthma Learning Health System dataset analysis yielded suggestive associations at rs35317849 near *CUL2* (HGNC:2552, p-value = 3.98 ×10^−7^; odds ratio = 2.14) and rs73846614 between *VGLL3* (HGNC:24327) and *CHMP2B* (HGNC:24537, p-value = 8.16×10^−7^; odds ratio = 3.48). These variants have not previously been associated with asthma or asthma outcomes. The suggestive associations at these loci highlight a need for additional studies that increase statistical power by integrating additional well characterized participants. Additionally, the results provide rationale to develop tools that incorporate genetic markers into models of asthma exacerbations and emergency care needs (**Figure 5A**).

**Figure 5.**
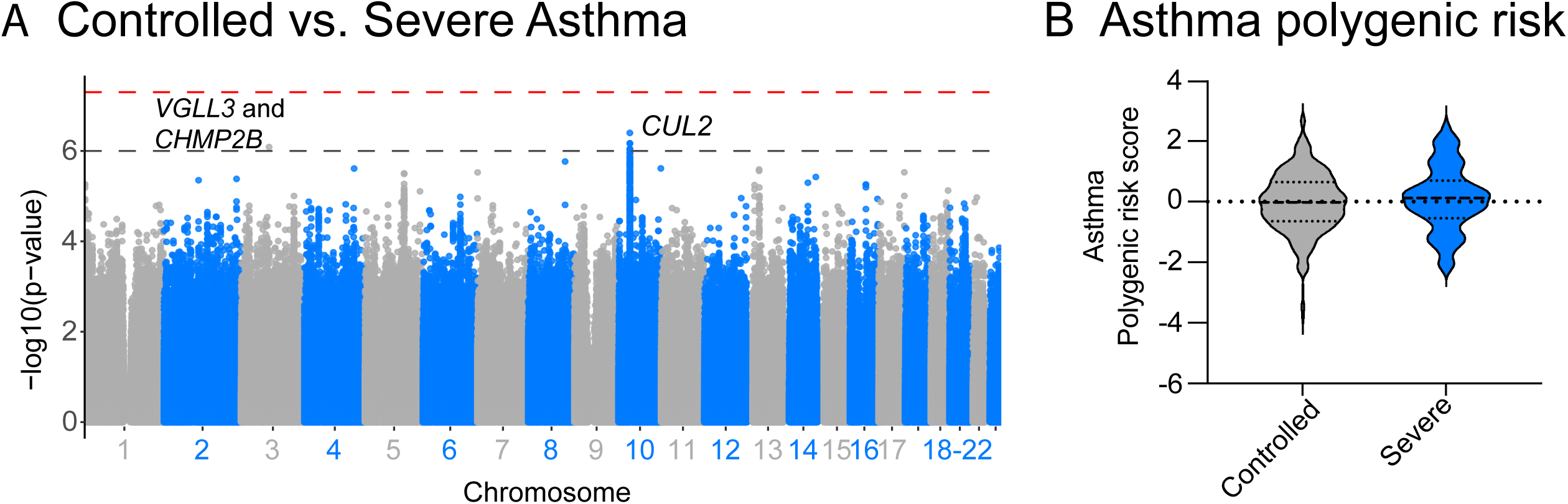
Partnerships to connect expert clinical phenotyping with high quality genetic data: acute asthma exacerbation. A. Individuals who were identified by clinicians as having controlled asthma and those with acute asthma exacerbations (labeled as severe asthma) were compared in a genome-wide association study. The genomic inflation for this analysis was 1.021. B. A polygenic risk score for asthma was used to quantify the cumulative burden of asthma genetic risk loci for each individual in the study^53^ (PRS002754, PGP000335). The difference in the distribution of polygenic risk was not statistically significant (one sided p-value of 0.108).

To further understand the role of combined genetic individual risk for individuals in this cohort, we calculated polygenic risk scores using a tool that was previously developed and validated across multiancestral asthma cohorts^53^: PRS002754, PGP000335. While not significantly different, the participants who required more acute care trended towards having a higher burden of asthma risk variants (one sided p-value of 0.109) (**Figure 5B**). Moreover, because we were able to provide a single datapoint that quantifies the cumulative genetic risk for each individual in this study, the clinical investigators were able to use this score to adjust for genetic risk when assessing other environmental and clinical factors. Together, these results highlight opportunities for integrating genetic research into precision medicine approaches for asthma management.

This sequencing resource highlights opportunities for discovery of genomic mechanisms by integrating genetic findings with genomic discoveries in cohorts with African, Puerto Rican, and Mexican ancestry. For example, of variants with significant or suggestive genetic association across the four genome wide association studies presented, most are also associated with allelic gene expression based on the Zendo expression quantitative trait loci (eQTL) database^54^ which identified ancestry-dependent eQTLs (**Supplemental Dataset 6**).

For Sickle cell disorder, genome-wide associated variants were associated with the expression of hemoglobin subunit gamma-1 (*HBG1*; HGNC:4831) and hemoglobin subunit gamma-2 (*HBG2*; HGNC:4832) in addition to members of the tripartite motif (TRIM) family, which are indued by interferons and play a role in immunological defenses ^55^. Variants reaching nominal Anxiety were associated with the expression of inositol polyphosphate multikinase (*IPMK*; HGNC:20739). IPMK regulates the mTOR pathway, with mTOR being important in mood regulation ^56^. Asthma associated variants were associated with genotype-dependent expression of three genes at the 17q12-21 locus: Gasdermin B (*GSDMB*), ORMDL Sphingolipid Biosynthesis Regulator 3 (O*RMDL3*; HGNC:16038), and Post-GPI Attachment to Proteins Phospholipase 3 (*PGAP3*; HGNC:23719). While GSDMB and ORMDL3 are often reported as important asthma genes, PGAP3 is also important, as it regulates human bronchila epithelial cell gene expression associated with asthma ^57^. From the Asthma Health Learning Network study, the associated haplotype on chromosome 10 is associated with cAMP-responsive element modulator (*CREM*; HGNC:2352) expression. CREM is a negative regulator of T helper 2 cytokine responses, which are central to allergic asthma.

## Discussion

In this study, we established a comprehensive sequencing and phenotyping resource that addresses the historical underrepresentation of individuals of African ancestry in genetic studies, particularly in pediatric disease research. By leveraging the Discover Together Biobank at CCHMC, we produced a rich dataset comprising deep genetic and phenotypic data from 15,684 children who self-identified as African American or Black. The study was made possible using low-coverage (3.3X) genome sequencing and an innovative imputation method to accurately call genotypes. We demonstrate that the resulting data are of high quality using heterozygous concordance of samples from the 1000 genomes project as well as samples from this cohort for which we had corresponding 30X genome sequencing data.

In this resource, we selected inclusion thresholds that maximized heterozygous concordance with control samples for which we also had high coverage genome wide sequencing data. The limitation of this approach is that it likely removed some high-quality variants. For example, the Hardy Weinberg Equilibrium threshold of p>0.0001 is especially conservative considering the gene flow assumptions of admixed and culturally heterogenous populations ^58^. This dataset is appropriate for the identification of common variants with allele frequencies greater than 1%. Additional individuals with genotype data will likely be needed to obtain sufficient statistical power to robustly identify genome-wide genetic associations with small to moderate genetic effects. For rare diseases with large effect size monogenic contributors, additional exome sequencing, would be needed to identify monogenic variants. Once identified, the common genotypes in this resource could be used to assess genetic modifiers of monogenic penetrance^59–62^.

We present a framework for case-control assessment using this cohort and show opportunities for discovery by presenting exemplary analyses. Our GWAS identified new and replicated previously known significant genetic associations for sickle cell disease, asthma, anxiety, and severe asthma - demonstrating the power of this resource to advance our understanding of the genetic underpinnings of pediatric diseases. This study is non-comprehensive and did not attempt to present all possible genetic analyses from this sequencing resource. While we did identify novel associations with anxiety and asthma as well as novel suggestive associations with asthma and asthma outcomes, these will also need to be replicated in subsequent studies

Our collaboration with a Community Advisory Committee has been instrumental in ensuring that our research is conducted ethically and inclusively. The committee’s insights guided our efforts to increase the representation of historically underrepresented populations in genomic studies and to develop community engagement strategies that promote transparency and trust. This partnership has also informed our decision to prioritize genetic studies of conditions that disproportionately impact Black and African American children, such as asthma and anxiety.

The genetic diversity observed in our cohort, of African ancestry ranging from 2% to 99%, highlights the potential to uncover ancestry-specific genetic variants that may contribute to disease risk. This range of admixture also creates an opportunity for admixture-based genetic studies that leverage the differences in local and global admixture to identify variants associated with phenotypes with ancestry-specific differences. These opportunities are particularly relevant for conditions such as sickle cell disease, anxiety, and asthma, each with morbidity which disproportionately affect Black and African American children. Gaining information from people of diverse ancestral backgrounds is also essential to fully capture the biological mechanisms underlying health and disease ^63^. For the first 20 years after the initial human genome was released, a European-centered “reference genome” has been the basis for genetic and genomic discovery. Seventy percent of the current GRCh38, for example, is based on one man of European ancestry. In this study we used a reference-genome-free approach that is not able to be used in all genetic and genomic studies. Expanding sequencing resources to include individuals of non-European ancestry also provides opportunities to better define and develop a human pangenome that is representative of global human genomic variation.

In this manuscript we do not present any family-based analyses. In future studies that use this resource, inclusion of related individuals, such as multi-generational parent-child dyads and siblings, will allow for the investigation of heritability and the identification of genetic factors that contribute to health outcomes within families. Additionally, the integration of electronic medical record data with genetic data enables the exploration of gene-environment interactions and the impact of socio-environmental factors on health outcomes.

Since this sequencing dataset was derived from Discover Together Biobank and made searchable via the Genomic Information Commons cohort builder, the dataset is well positioned to be easily accessible via a transparent, equitable, and tracked request system. The review process is built around the Genomic Information Commons sample request tool, with review procedures following the Discover Together Biobank protocols in place for institutional samples and data. Researchers at institutions that are part of the Genomic Information Commons can request data for analyses. Additionally, researchers who are not at institutions that are part of the Genomic Information Commons can find collaborators at a Genomic Information Commons -member institution to collaborate on a data request.

In conclusion, the unique sequencing resource we developed represents a significant step towards addressing the disparities in genomic research and advancing precision medicine for all. By providing high-quality genetic and phenotypic data, this resource has the potential to drive discoveries that improve health outcomes for children of African ancestry. Future research should continue to leverage this resource to explore the genetic basis of pediatric diseases and to develop targeted interventions that promote health equity.

## Data Availability

The data described in this manuscript have been deposited in the GIC (https://www.genomicinformationcommons.org/).

## Acknowledgements

The authors would like to acknowledge the CCHMC Genomics Sequencing Facility (RRID:SCR_022630). This study used samples, data, and/or services from Cincinnati Children’s Discover Together Biobank [RRID: SCR_022632]. We thank the Discover Together Biobank for support of this study and the participants and their families, whose help and participation made this work possible. Schematic figures were made in BioRender.

## Author Contributions

Conceptualization: LCK, IC, LM, LBH, KMK, ; Data curation: LCK, SR, MT, LPL, AC, BN, IC, KMK; Formal analysis: LCK, KMK, BN; Funding acquisition: LM, JBH; Visualization: LCK, KMK; Writing-original draft: LCK and KMK; Writing-review & editing: LCK, SR, MT, LPL, SE, MH, JM, AC, LT, YH, CF, AS, TA, MM, AB, MS, NU, SF, BC, MB, MA, TM, BN, MWP, JRS, RTA, DS, JP, TG, CP, LJM, IC, LM, JH, KMK.

## Ethics Declaration

The CCHMC Institutional Review Board (IRB) reviewed and approved this study. As detailed in the methods section, samples were obtained from the Discover Together Biobank after approval from their sample use committee. Such participants consented for their samples and data to be used for future research. The Discover Together Biobank Biospecimen and data Access Committee reviewed and approved for such consented samples to be used for this project.

---

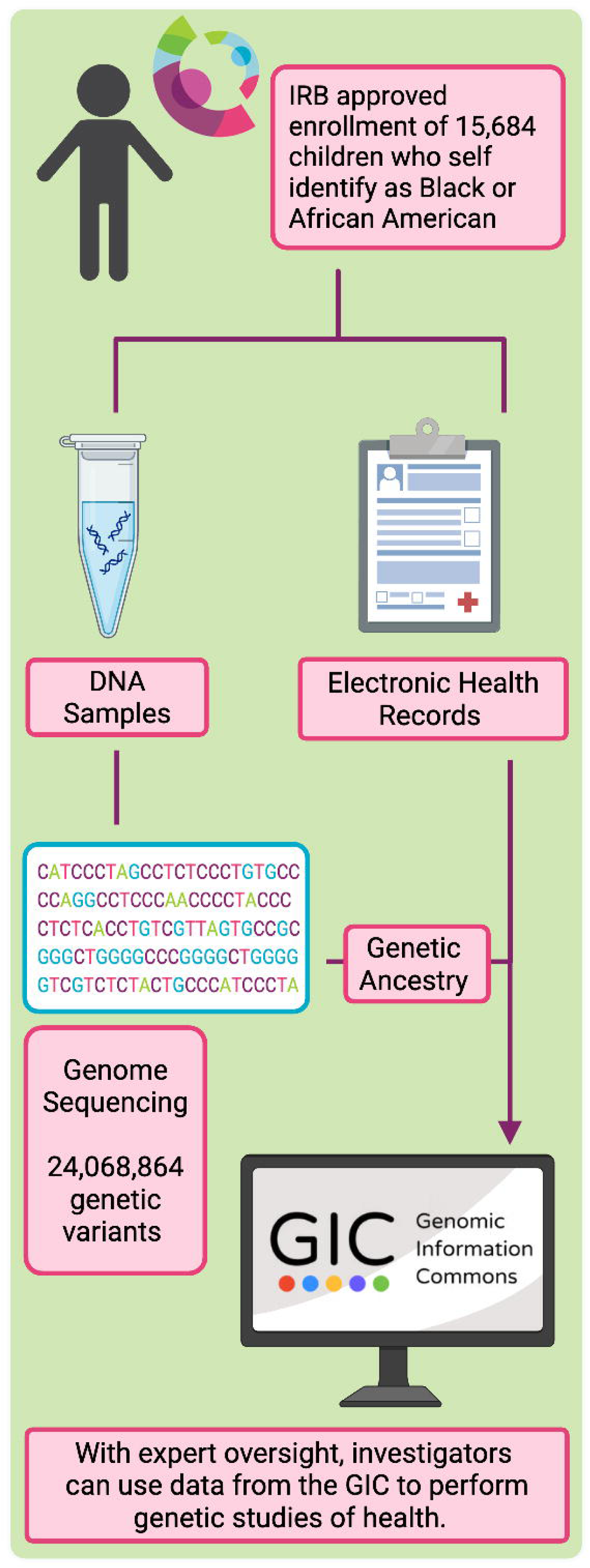

**Supplemental Figure 1.**
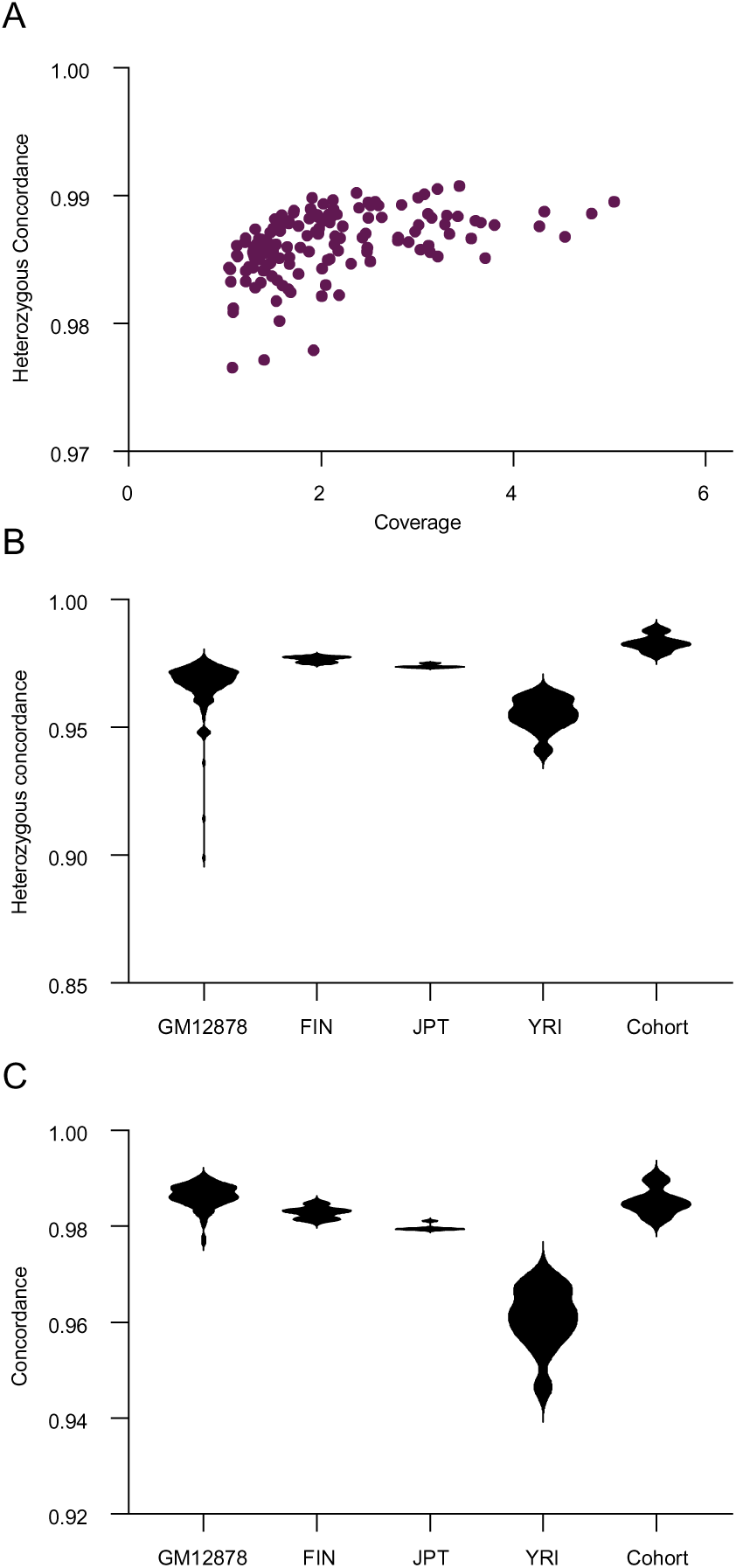
Quality assessment of sequencing data. Heterozygous concordance refers to the rate at which a variant is correctly identified as heterozygous (having one copy of each allele) across sequencing studies. A. Heterozygous concordance (y-axis) of GM12878 for each sequencing run of this study was assessed using corresponding 30X sequencing and is shown in the context of genomic coverage (x-axis). Heterozygous concordance (B) and full concordance (C) is presented for each control sample from which there was corresponding 30X sequencing data (see Methods).

**Supplemental Figure 2.**
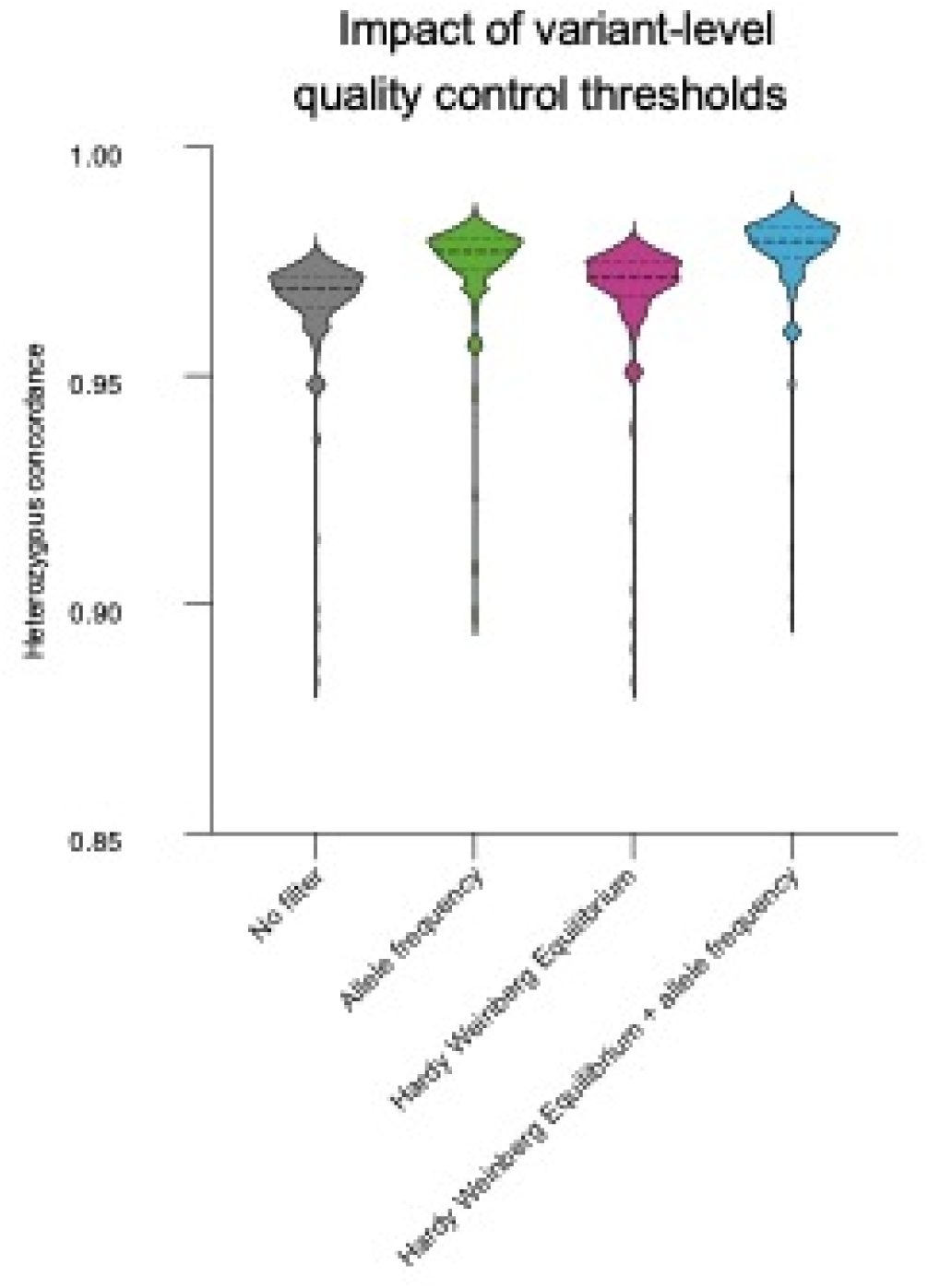
Impact of variant-level quality control thresholds. Heterozygous concordance for low-pass sequencing of GM12878 is presented with no filters, with a 1% allele frequency filter, with a Hardy-Weinberg equilibrium threshold of 0.0001, and with both allele frequency and Hardy-Weinberg filters.

**Supplemental Figure 3.**
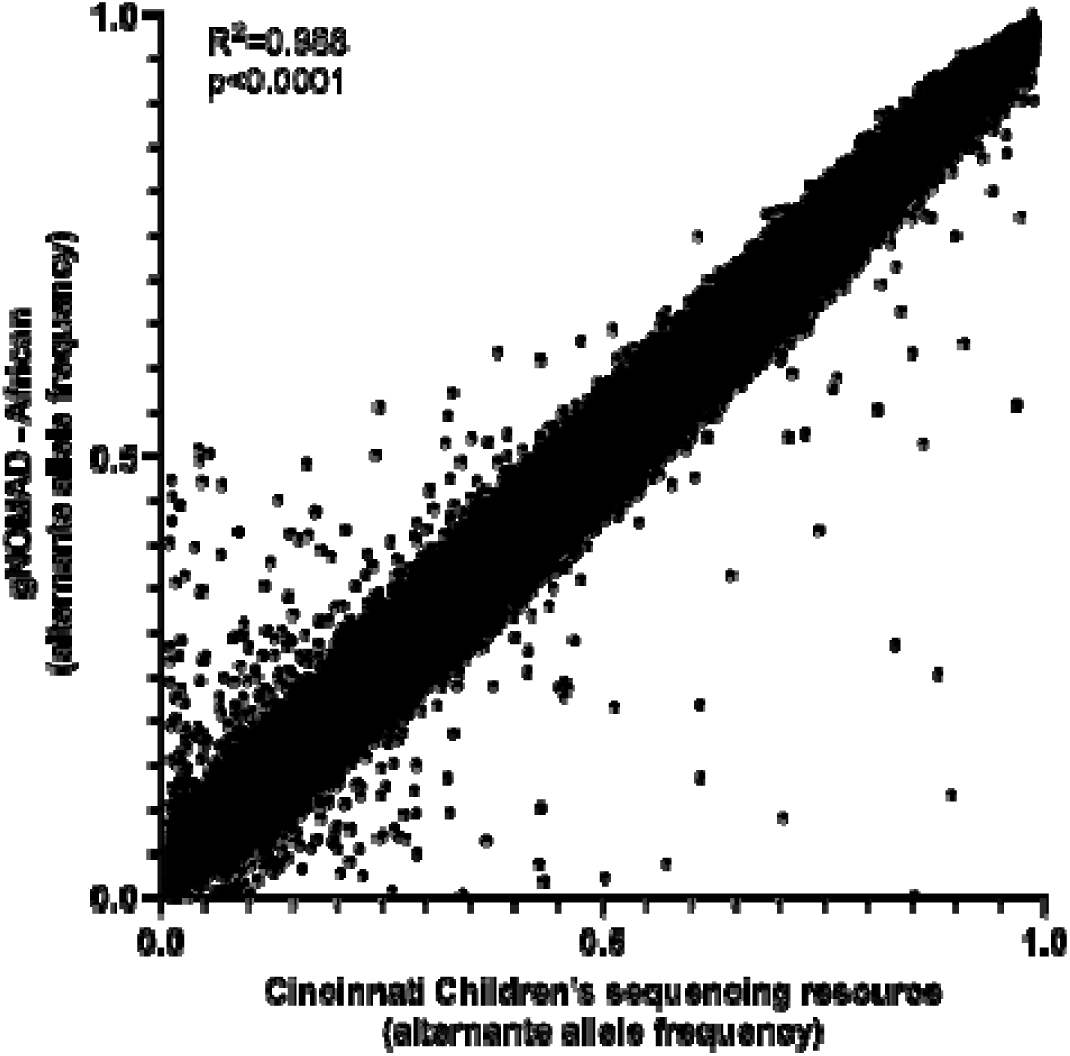
Correlation of alternative allele frequencies in this sequencing resource compared to the population of African ancestry in gnomADv4.0. The correlation of allele frequencies was calculated using the Person method using 13,648 unrelated individuals. In this graph, allele frequencies from a random 100,000 variants are shown. The full set of allele frequencies for the entire cohort is available in **Supplemental Dataset 1**.

**Supplemental Figure 4.**
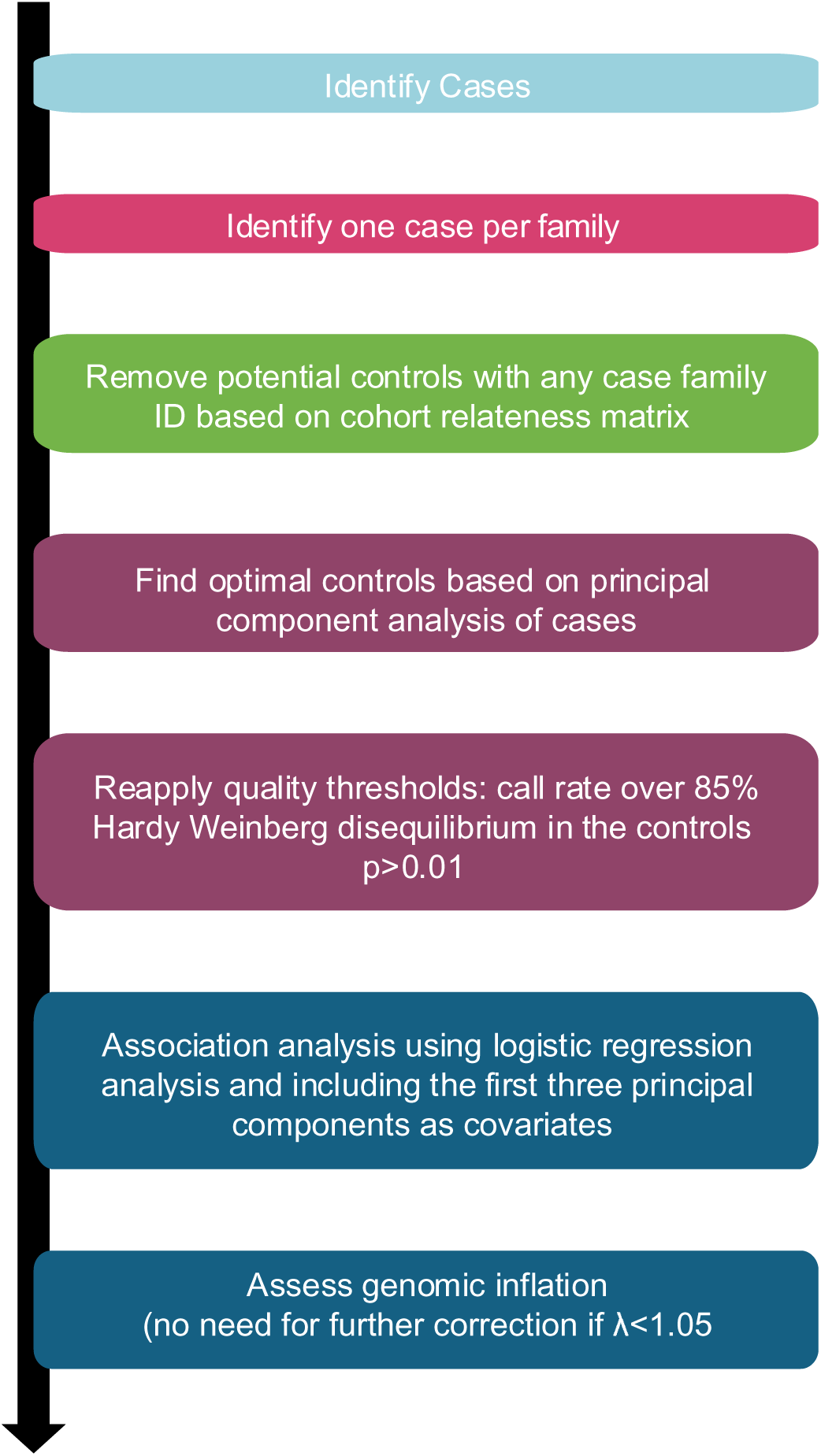
General recommended analytical strategy for case control analyses using this cohort.

