## Supplementary material for "Sequencing and health data resource of children of African ancestry": Table 1

**Table 1. Relatedness of cohort out of 15,684 individuals**. A full list of individuals in our cohort and their relatedness to other participants in the cohort can be found in Supplemental Dataset 2.

| **Relation** | **Count** |
| --- | --- |
| Monozygotic twins | 41 |
| Parent - Offspring | 98 |
| Siblings | 1,050 |
| 2nd degree | 134 |
