## Supplementary material for "Sequencing and health data resource of children of African ancestry": Table 2

**Table 2. Top 20 International Classification of Diseases (ICD)-10 codes associated with cohort**

| **Number** | **ICD-10**  **code** | **Description** |
| --- | --- | --- |
| 2,916 | J06.9 | Acute upper respiratory infection, unspecified |
| 2,446 | R50.9 | Fever, unspecified |
| 2,137 | R05 | Cough |
| 2,033 | J45.909 | Unspecified asthma, uncomplicated |
| 1,963 | R11.10 | Vomiting, unspecified |
| 1,915 | K59.00 | Constipation, unspecified |
| 1,813 | B34.9 | Viral infection, unspecified |
| 1,769 | R10.9 | Unspecified abdominal pain |
| 1,751 | Z23 | Encounter for immunization |
| 1,747 | J02.9 | Acute pharyngitis, unspecified |
| 1,745 | Z20.822 | Contact with and (suspect exposure to covid-19) |
| 1,548 | R51 | Headache |
| 1,326 | R09.81 | Nasal congestion |
| 1,305 | Z32.02 | Encounter for pregnancy test, result negative |
| 1,257 | B97.89 | Other viral agents as the cause of diseases classified elsewhere |
| 1,235 | E66.9 | Obesity, unspecified |
| 1,220 | Z53.21 | Procedure and treatment not carried out due to patient leaving prior to being seen by health care provider |
| 1,204 | F90.9 | Attention-deficit hyperactivity disorder, unspecified type |
| 1,155 | R19.7 | Diarrhea, unspecified |
| 1,086 | L30.9 | Dermatitis, unspecified |
