## Supplementary material for "Sequencing and health data resource of children of African ancestry": Table 3

**Table 3. Top 20 Current Procedural Terminology (CPT) and Healthcare Common Procedure Coding System (HCPCS) Codes associated with cohort**

| **Number** | **CPT or HCPCS code** | **Description** |
| --- | --- | --- |
| 14,078 | 99213 | Established patient office visit, 20-29 minutes |
| 13,444 | 36415 | Routine venipuncture |
| 11,569 | 85025 | Comprehensive blood count (CBC) with automated differential count |
| 11,216 | 99283 | Emergency department visits that require moderate complexity medical decision making |
| 10,231 | 99214 | Office or outpatient visit for an established patient |
| 10,058 | 90471 | The administration of a single vaccine or combination vaccine by injection |
| 9,946 | 99212 | An evaluation and management (E/M) visit for an established patient |
| 9,572 | 94760 | A single noninvasive pulse oximetry measurement |
| 8,790 | 99284 | An emergency department (ED) visit for a patient with a high-severity problem |
| 8,176 | 99211 | An office or outpatient visit for an established patient that may not require a physician |
| 7,991 | 84443 | Laboratory test that measures thyroid-stimulating hormone (TSH) levels |
| 7,831 | 85027 | Complete blood count (CBC) that doesn't include a white blood cell differential |
| 7,695 | 90472 | Additional vaccine given after the first vaccine |
| 7,534 | 80061 | Lipid panel |
| 7,418 | 99282 | Emergency department visit for a new or established patient. |
| 7,405 | 1445253 | Closed Reduction, Splinting Right Thumb |
| 7,354 | 81001 | Urinalysis performed with a dipstick or tablet reagent and microscopy |
| 7,182 | 506675 | Hypertrophy of Tonsils and Adenoids |
| 7,150 | 1435700 | Spirometry Pre & Post Bronchodilators |
| 7,123 | 1481540 | Tonsillectomy and Adenoidectomy |
