## Supplementary material for "Sequencing and health data resource of children of African ancestry": Table 4

**Table 4. Top 20 Medications associated with cohort**

| **Number** | **Medicine Name** |
| --- | --- |
| 8,692 | Ibuprofen 100 mg/5mL oral suspension |
| 8,678 | Normal saline flush for medications |
| 8,598 | Acetaminophen 160 mg/5mL oral suspension |
| 7,962 | Intravenous fluids |
| 7,094 | Sodium chloride 0.9% intravenous / injectable solution |
| 6,601 | Influenza virus vaccine split intramuscular suspension |
| 6,502 | Sodium chloride 0.9% injectable solution |
| 6,444 | Acetaminophen 325 mg oral tabs |
| 6,303 | Ibuprofen 200 mg oral tabs |
| 6,049 | Ondansetron HCL 4 mg/2mL injectable solution |
| 5,526 | Hepatitis A vaccine 720 el u/0.5mL intramuscular suspension |
| 5,303 | Morphine sulfate (preservative-free) 1 mg/mL injectable solution |
| 5,273 | Sodium chloride 0.9 % intravenous bolus infusion |
| 5,158 | Albuterol sulfate HFA 108 (90 base) mcg/act inhaled Aerosol |
| 4,935 | Albuterol sulfate (2.5 mg/3mL) 0.083% inhaled nebulizer |
| 4,892 | Amoxicillin 400 mg/5mL oral suspension |
| 4,891 | Lactated ringers intravenous solution |
| 4,694 | Fentanyl citrate 0.05 mg/mL injectable solution |
| 4,569 | Dexamethasone sodium phosphate 4 mg/mL injectable solution |
| 4,545 | Acetaminophen 10 mg/mL intravenous solution |
