## Supplementary material for "Sequencing and health data resource of children of African ancestry": Table 5

**Table 5. Demographic and genomic characteristics of presented association studies.** After identifying controls that were matched to cases based on admixture and removing first and second-degree relatives, genome wide association analyses were performed in PLINK1.9 using a logistic regression that adjusted for admixture estimates and genomic inflation.

| **Phenotype** | **Number of cases** | **Number of controls** | **Percent female cases** | **Percent female controls** | **Average African admixture cases** | **Average African admixture controls** | **Genomic inflation** |
| --- | --- | --- | --- | --- | --- | --- | --- |
| Sickle cell disease | 753 | 4518 | 46.2% | 54.7% | 0.182 | 0.192 | 1.033 |
| Anxiety | 3740 | 7480 | 55.6% | 52.7% | 0.245 | 0.238 | 1.048 |
| Asthma | 3740 | 7239 | 47.7% | 56.6% | 0.227 | 0.236 | 1.042 |
| Severe vs. controlled asthma | 99 | 393 | 44.4% | 44.5% | 0.221 | 0.204 | 1.021 |
