## Supplemental Note for "Sequencing and health data resource of children of African ancestry"

**Suggested analytical protocol for case – control studies using this cohort**

Based upon the range of admixture and relatedness in this cohort, we developed a case-control analytical framework for performing genome-wide association studies using individuals in this sequencing resource (**Supplemental Figure 4**). First, it is best practice to identify cases and controls based on validated electronic medical record-based phenotyping algorithms^44^. When such algorithms are not available, care should be taken to develop inclusion and exclusion criteria for cases and controls in collaboration with physicians who are familiar with electronic medical record-based coding of the health outcome of interest^44^. Next, one case should be chosen from each family^45^. Controls can be identified using an algorithm with inclusion and exclusion criteria that are appropriate for the phenotype being studied. Controls should be identified from those who are not related to cases. The controls identified for a study should then be matched to cases based on the principal component analysis^40^. Before initiating association testing, call rate and Hardy Weinberg disequilibrium quality thresholds should be reapplied. Case control analyses should use logistic regression and include informative principal components of each individual as covariates, as informed by an analysis of the scree plot^46^. Likewise, continuous variable analysis should use linear regression with principal components as covariates^46^. Using this framework, the genomic inflation should be assessed, with an optimal value below 1.05. Values over 1.05 suggest the need for more careful matching of controls to cases^47^. We have found that one way to improve the matching of controls to cases using principal component measurements is to decrease the requested ratio of controls to cases.

**Supplemental Methods**

**Quantifying DNA and preparing samples for sequencing**

DNA was obtained from the Discover Together Biobank at a requested concentration of 50 ng/μL via UV absorbance spectroscopy. SYBR Green fluorescence was used to measure DNA concentration. It is important to utilize the same amount of DNA for each sample to generate uniform DNA sequencing coverage. To generate accurate DNA stocks, we performed three serial dilutions (25 ng/μL, 10 ng/μL and 2.5 ng/μL) using SYBR Green fluorescence to measure DNA concentration after each dilution. The final DNA concentration for all samples was between 2.4 and 2.6 ng/μL.

DNA from a subject included in HapMap and the 1000 genomes project, GM12878, was included within each sequencing batch of this study to allow the study team to confirm appropriate sample tracking. The inclusion of additional control samples also allowed us to assess concordant genotype calls in this study compared to prior 30X whole genome sequencing studies. Most genetic variants in a person are homozygous for the reference allele, so using global genotype concordance or homozygous concordance can result in a superficially enhanced assessment of quality^32^. Heterozygous calls are more susceptible to errors in sequencing data and the concordance of heterozygous variance is less susceptible to bias because of overrepresentation of the reference allele. “Heterozygous concordance” refers to the rate at which a variant is correctly identified as heterozygous (having one copy of each allele) across two or more sequencing studies^32, 33^.

Before applying quality thresholds, GM12878 heterozygous concordance ranged from 89.7% to 98.7%. Heterozygous concordance was correlated with up to a genomic coverage of 2 after which it did not meaningfully improve further (**Supplemental Figure 1**). Samples with coverage less than 1 had an average heterozygous concordance of 91.0%, so we set the genomic coverage threshold for our study at 1X, resulting in an average heterozygous concordance of 96.6%. Adding an allele frequency threshold of 1% and a Hardy-Weinberg equilibrium threshold of 0.0001 further increased heterozygous concordance to 97.7% (**Supplemental Figure 2**). Samples from 1000 genomes of other ancestries were also included in various sequencing batches, with a heterozygous coverage across all assessed 1000 genomes controls of 97.53% (**Supplemental Figure 1**). A batch of 11 samples from the current resource were assessed with 30X whole genome sequencing, and the heterozygous concordance of these samples was 98.3% with a total concordance of 98.5% (**Supplemental Figure 1B and 1C**).

**Control Samples**

Each 96 well plate for this project included one or two aliquots of DNA from the human lymphoblastoid cell line GM12878 (ATCC Manassas, Virginia) that were randomly plated into different wells on each plate. These samples served as a control for sequencing quality and sample tracking. Concordance between control samples was calculated using the software tool SnpSift^34^.

**Library Preparation**

Two methods were used to generate DNA sequencing libraries for this project. The first 384 samples were processed with the Riptide High Throughput Rapid Library Prep (HT-RLP) kit from iGenomiX San Fransico, CA (now owned by TWIST Bioscience Corporation). All remaining samples were processed with the SeqWell (Beverly, MA) plexWell™ 96 NGS multiplexed library generation kit for Illumina sequencing. All libraries were constructed using the manufacturers’ recommended procedures.

**Next generation sequencing and analysis**

Illumina (San Diego, CA) short read sequencing was performed by the CCHMC Genomics DNA Sequencing Facility utilizing an Illumina NovaSeq 6000 instrument and Illumina S4 sequencing cells. All sequencing was performed following Illumina’s standard sequencing protocol.

All computational processing of Illumina DNA sequencing data was performed on the CCHMC computational cluster. The CCHMC Genomics DNA Sequencing Facility provided individual fastq files for each sample after library sequencing. Each sample was aligned to the human genome build GRCh38 using BWA (version 0.7.17-r1188)^35^ using the following options: bwa mem -t 8 -B 4 -O 6 -E 1 -M -R. Duplicate reads were identified and removed using the picard software package (<https://broadinstitute.github.io/picard/>) with the command picard MarkDuplicates REMOVE_DUPLICATES=true OPTICAL_DUPLICATE_PIXEL_DISTANCE=2500. Quality control measurements were obtained for each bam file using the software package Bamtools with the “stats” command^36^.

Average autosome coverage and chromosome X coverage were calculated using read counts generated by the command samtools idxstats^37^. Sex for each sample was determined by calculating the average autosome coverage/average Chromosome X coverage. Ratios between 0.4 and 0.6 were considered male and ratios between 0.9 and 1.1 were considered female. Samples outside these ranges were considered indeterminant. All samples with an average autosome coverage less than 1.0 and an indeterminant sex determination or mismatched estimated sex and reported sex were dropped from the study. Genotypes were called using the software program STITCH^30^ on batches of samples ranging from 96 to 384 per run. In addition to the biorepository samples, each genotype calling batch included BAM files from 997 individuals obtained from the 1000 Genomes Project, each down-sampled to an average of 1x sequencing coverage.
